# Trans Sodium Crocetinate (TSC) to Improve Oxygenation in COVID-19

**DOI:** 10.1101/2021.10.08.21264719

**Authors:** Anca Streinu-Cercel, Oana Săndulescu, Victor Daniel Miron, Alina-Alexandra Oană, Maria Magdalena Motoi, Christopher D. Galloway, Adrian Streinu-Cercel

## Abstract

**Background:** Trans Sodium Crocetinate (TSC) is a bipolar synthetic carotenoid under development as a drug to enhance oxygenation to hypoxic tissue in addition to standard of care. TSC acts via a novel mechanism of action, improving the diffusivity of oxygen in blood plasma. Thus, it is based on physical-chemical principles, unlike most drugs which are based on biochemistry-based mechanisms. We explored the use of escalating doses and multiple daily dosing of TSC as a potential therapeutic for patients suffering from hypoxemia due to SARS-CoV-2 infection.

**Methods:** Individuals ≥18 years who were hospitalized with confirmed SARS-CoV-2 infection and hypoxemia, defined as SpO_2_ < 94% on room air or requiring supplemental oxygen, WHO ordinal scale 3 through 7 (exclusive of Extra Corporeal Membrane Oxygenation [ECMO]) were enrolled in cohorts of six subjects, each of whom received the same dose (0.25, 0.5, 1.0, or 1.5 mg/kg) of TSC via intravenous bolus every 6 hours in addition to standard of care (SOC).

This report describes the safety and efficacy results from the lead-in phase of the study and the population pharmacokinetics (PK) analyses. Safety was assessed as the number of serious adverse events and dose-limiting toxicities (DLTs) observed with each dose. Several efficacy parameters were examined in the lead-in phase and descriptive statistics of efficacy parameters are provided. No formal statistical analyses were performed. The population PK analyses were based on previous analyses and examination of the concentration profiles, and two-compartment linear pharmacokinetic models were evaluated and validated. Covariates, including body size, age, sex, organ function, and dose level, were evaluated for inclusion into the model.

**Results:** TSC was well tolerated. There were no treatment emergent adverse events (TEAEs) reported. There were 2 serious adverse events (SAEs) reported during the study, neither were considered treatment-related. A total of 24 (96%) subjects survived. One subject (4.0%) died during the study as a result of an SAE (respiratory failure), and that event was determined to be due to COVID-19 complications and not related to study drug.

There was an observed reduction in the time to improvement in WHO Ordinal Scale with increasing dose. The median time to 1-point reduction in subjects receiving 0.25 mg/kg was 11.5 days versus 7.5 days in the 1.5 mg/kg treatment cohort. The overall range across all doses was 1 day to 28 days. A total of 36.0% of subjects had a 1-point improvement in WHO Ordinal Scale to Day 7. The 1.5 mg/kg dose resulted in observed superior outcomes for multiple secondary clinical outcomes: time to 1-point WHO Ordinal Score improvement through Day 29/discharge, 1-point improvement by Day 7, days to return to room air, and hospital length of stay.

The PK results showed that the two-compartment model fit the data well. Clearance decreased with increasing dose level and there was no evidence that clearance was affected by covariates other than dose level.

**Conclusions:** These findings suggest that TSC administration every 6 hours at doses up to 1.5 mg/kg for up to 15 days is safe and well tolerated with predictable pharmacokinetics and demonstrated an observed clinical benefit in the treatment of COVID-19-related hypoxemia.

(ClinicalTrials.gov number, NCT04573322)

## INTRODUCTION

The novel coronavirus, designated as SARS-CoV-2, and the disease caused by this virus, COVID-19, is still prevalent in many areas of the world, despite widespread vaccination. Globally, as of 17 September 2021, there have been 226,844,344 confirmed cases of COVID-19, including 4,666,334 deaths, reported to WHO.^1^ Current acute treatment options continue to evolve and and may include the use of remdesivir, an antiviral, anti-inflammatory medications; as well well as monoclonal antibody infusions. These have shown some efficacy particularly when administered early in the infection and can all be part of multi-modal therapy for affected patients. Remdesivir has known safety issues in patients with renal dysfunction due to its formulation with sulfobutylether beta-cyclodextrin sodium^2^.

Patients suffering from moderate to severe COVID-19, with pneumonia, could rapidly progress to acute respiratory distress syndrome (ARDS).^3,4^ These patients can often require hospitalization for oxygen supplementation as well as admission to an intensive care unit where mechanical ventilatory (MV) support and possibly extra-corporeal membrane oxygenation (ECMO) or even lung transplant may be needed. COVID-19 leading to severe ARDS is associated with a high degree of morbidity and mortality. Prior to developing ARDS, patients with COVID-19 may experience a period of so-called silent hypoxemia, consisting of observable hypoxemia by oxygen saturation (SpO_2_) measurements, but showing minimal outward signs of respiratory distress.^5^ Impairment of cellular oxygenation is a key factor in the pathophysiology of COVID-19. Endothelial damage can contribute to ventilation-perfusion mismatch leading to hypoxemia,^6^ and in turn severe hypoxemia may lead to multiple organ failure and death.^7^

Trans sodium crocetinate (TSC) is a novel drug developed to enhance oxygenation to hypoxic tissue. This study evaluated the safety, pharmacokinetics, and efficacy of TSC to improve oxygenation and clinical outcomes in hospitalized hypoxemic patients infected with SARS-CoV-2.

## METHODS

### Trial Design

The study was designed to have two phases: an open-label lead-in phase and a randomized placebo-controlled phase. The results described herein are from the lead-in phase of the study. This was an open-label, pharmacokinetic, pharmacodynamic, multiple daily, ascending dose, safety and tolerability study in SARS-CoV-2 infected patients with hypoxemia requiring hospitalization (ClinicalTrials.gov number, NCT04573322). The trial was conducted at a single study center in Romania, at The National Institute for Infectious Disease.

The lead-in study enrolled 25 subjects. Eligible subjects received an IV bolus of TSC four times daily (every six hours) for up to 15 days (minimum of 5 days). This study utilized an ascending dose scheme (6 patients per dose cohort), starting with a dose of 0.25 mg/kg.

The protocol is available upon request.

### Ethics Statement

The study was conducted in accordance with the general ethical principles as enunciated in the Declaration of Helsinki (1996) and in conformance with International Conference on Harmonisation (ICH) Good Clinical Practice (GCP) guidance and applicable Health Authority and local requirements regarding Institutional Review Boards, Independent Ethics Committees, informed consent, and other statutes or regulations related to the rights and welfare of human subjects participating in biomedical research.

### Study Participants

The study population consisted of hospitalized adult (≥18 years of age) patients with confirmed SARS-CoV-2 infection and hypoxemia, defined as SpO_2_ < 94% on room air or requiring supplemental oxygen, WHO ordinal scale 3-7. Full inclusion and exclusion criteria are provided in the Appendix A.

### Intervention and Assessments

Beginning on Study Day 1, eligible subjects received an IV bolus of TSC four times daily (every six hours) for up to 15 days (minimum of 5 days). Each TSC dose was administered via IV bolus to 6 subjects per dose level (0.25, 0.5, 1, or 1.5 mg/kg). One subject from the 1.0 mg/kg dose group received one dose of TSC and withdrew therefore an additional patient was enrolled in this cohort. This study utilized a 3 + 3 design with an ascending dose scheme. The first group of 3 subjects received TSC at 0.25 mg/kg. If no dose limiting toxicities (DLT) had occurred, an additional 3 subjects were to receive 0.25 mg/kg TSC. If there were <33% observed toxicities among 6 subjects, then another 3 subjects were to receive the next dose level up (0.5 mg/kg) and so forth. An Independent Safety Monitoring Committee (SMC) was established to review the safety data of a given cohort and approved or disapproved enrollment of the next cohort. At the completion of the lead-in portion of the study, the SMC examined resultant safety data and blood oxygenation data for all subjects and determined all doses were safe and well tolerated with observation of improved clinical outcomes at the highest doses tested.

Key assessments included weekly SARS-CoV-2 reverse transcription polymerase chain reaction (RT-PCR) analysis, WHO 9-point Ordinal Scale assessment (daily, first assessment of the day), Glasgow Coma Score (GCS; daily), Sequential Organ Failure Assessment (SOFA; daily), and blood oxygenation was measured via recorded continuous pulse oximetry and the S:F ratio was calculated. Measures of clinical support were assessed and recorded during hospitalization: oxygen requirement, prone positioning, use of remdesivir or other antivirals, use of corticosteroids, non-invasive mechanical ventilation (via mask), mechanical ventilator requirement (via endotracheal tube or tracheostomy), and Extra Corporeal Membrane Oxygenation (ECMO) requirement.

The pharmacokinetics of TSC in blood were analyzed. A dataset suitable for a population pharmacokinetic analysis was assembled. The analyses were conducted with NONMEM software. Based on previous analyses and examination of the concentration profiles, the data were fit to a two-compartment linear pharmacokinetic model. Covariates, including body size, age, sex, organ function, and dose level, were evaluated for inclusion into the model. Model validation included a visual predictive check, a bootstrap analysis, and likelihood profiles for two parameters.

### Endpoints

This report describes the results of the lead-in period which was designed to assess safety and PK. Several efficacy parameters were examined in the lead-in phase, but no statistical analyses were performed. Descriptive statistics of efficacy parameters are provided.

The primary efficacy endpoint was the time to recovery through Day 28, defined as time to achieve (and maintain through Day 28) a WHO ordinal COVID-19 severity scale score of 1, 2 or 3 with a minimum 1-point improvement from baseline.

The secondary efficacy endpoints included changes in WHO ordinal severity scale, oxygenation parameters (including oxygen saturation to fraction of inspired oxygen ratio (SpO_2_/FiO_2_), ventilator-free days, and the time to return to room air), and length of hospitalization.

Safety endpoints included the incidence and severity of adverse events.

The appropriate pharmacokinetic model was selected based on several criteria: graphics displaying ratios of observed-to-population predicted values vs. time and observed vs. population predicted values were examined. A two-compartment linear model was evaluated initially. This was followed by evaluating whether body size should be incorporated into the model, then, the role of other covariates was evaluated.

### Statistical Analysis

No formal statistical analyses were performed. Descriptive statistics were provided for efficacy and safety endpoints.

## RESULTS

### Demographics and Baseline Characteristics

Thirty-one subjects were screened, and 25 subjects were enrolled in this study. The median age across all dose groups was 55 years and the majority of subjects (76%) were less than 65 years old. A total of 52% were female and all subjects were White. All subjects tested positive for SARS-CoV-2 infection.

Demographics and baseline characteristics for all subjects are presented in Table S 1.

### Efficacy

#### Time to recovery through Day 28

There was an observed reduction in the time to improvement in WHO Ordinal Scale with increasing dose. The median time to 1-point reduction in subjects receiving 0.25 mg/kg was 11.5 days versus 7.5 days in the 1.5 mg/kg treatment cohort. The overall range was 1 day to 28 days (Table S 2, Figure S 1).

A total of 9 (36%) subjects had a 1-point improvement in WHO Ordinal Scale to Day 7. One subject (4%) had a 1-point decline. No changes were seen in 48% of subjects (Table S 3).

SpO_2_/FiO_2_ is a marker for peripheral blood saturation of oxygen to fraction of inspired oxygen ratio and was measured for all patients prior to and at several timepoints after each dose was administered. The 1.0 and 1.5 mg/kg dose groups showed improvement in median pre-dose S/F ratio at Day 5 and later, not seen for 0.25 mg/kg and 0.50 mg/kg.

The median time required for a subject to return to room air was 7.5 days, ranging from 0.5 to 52 days. There appeared to be an effect with dose on this parameter, the median number of days for the 1.5 mg/kg group was 6.5 with a range of 0.5 to 6.5 days (Table S 4).

The length of hospitalization was assessed and the median length of stay across all dose groups was 8 days and overall range was between 2 to 14 days.

Overall, the 1.5 mg/kg dose resulted in observed superior outcomes for multiple secondary clinical outcomes: time to 1-point WHO Ordinal Score improvement through Day 29/discharge, 1-point improvement by Day 7, days to return to room air, and hospital length of stay, with only one death (96% survival observed).

Baseline (pre-dose) SpO_2_ ranged from 90.0 to 99.0 Day 1 and ranged from 92.0 to 100.0 on Day 10. Baseline FiO_2_ ranged from 0.28 to 0.75 Day 1 and ranged from 0.21 to 0.75 on Day 10. The median baseline ratio of SpO_2_/FiO_2_ was 217.80 on Day 1 and was 324.25 on Day 10. The change in median SpO_2_/FiO_2_ over time is presented in Table S 8 and graphically in Figure S 3, showing the improvement over time which was most notable at the highest doses of TSC studied.

### Safety

TSC was well tolerated. There were no TEAEs considered related to TSC. There were 2 subjects reporting SAEs, 1 death, and 1 TEAE leading to study drug discontinuation (Table S 5). No DLTs were reported at any dose level tested. A total of 14 (56%) subjects reported TEAEs. Of these, 5 (20%) subjects reported Grade 3 or 4 TEAEs. TEAEs from the SOC of Respiratory, Thoracic and Mediastinal Disorders (9 [36.0%] subjects) and Infections and Infestations (8 [32%]) were reported most frequently. Gastrointestinal disorders and Metabolism and Nutrition disorders were reported in 7 (28.0%) subjects, each. Most TEAEs were reported in 1 subject each. There were no clinically significant changes in laboratory assessments, vital signs, physical findings, or ECG parameters.

### Pharmacokinetics

Twenty-four subjects with 144 measurable timepoints were included in the PK analyses. One of the subjects was excluded from the analyses due to a PK profile that was inconsistent with a PK profile following an IV bolus dose. In addition, there were 11 trough samples with results that were much higher than expected, and likely taken after IV bolus rather than prior to the dose. These outliers were removed from the mean plasma concentration and the PK analysis. All pre-dose samples of the first dose were below the limit of quantitation (10 ng/mL) and were set to zero in the PK analysis.

The pharmacokinetics of TSC appear to be nonlinear saturable after IV doses of 0.25 to 1.5 mg/kg in patients with SARS-COV-2 infection (Figure S 2). Mean CL decreased from 5.38 mL/min/kg at 0.25 mg/kg to 1.62 mL/min/kg at 1.5 mg/kg. The mean half-life of TSC increased from 0.53 hours to 1.07 hours as dose increased from 0.25 mg/kg to 1.5 mg/kg. There was no significant accumulation of TSC after multiple IV administration of TSC every 6 hours (Table S 7).

Population pharmacokinetic analyses showed the two-compartment model fit the data well. Although graphics suggested that central compartment volume decreased with weight, models that incorporated an effect of weight or lean body mass on central compartment led to a consistent bias in the relationship between clearance and weight. Therefore, a model with no adjustment for body size was adopted. Increasing dose was associated with a decrease in clearance, distribution clearance, and volume of the peripheral compartment. There was no evidence that clearance was affected by concomitant medications, hepatic or renal laboratory variances, or other covariates beyond dose level.

## DISCUSSION

COVID-19 is a respiratory disease caused by a novel coronavirus (SARS-CoV-2) associated with substantial morbidity and mortality. This Phase 1/2 study was conducted in subjects infected with SARS-CoV-2 and was designed to evaluate the safety, pharmacokinetics, and efficacy of a novel agent, TSC, to enhance oxygenation in hospitalized, hypoxemic SARS-CoV-2 infected patients as an adjunct to standard of care.

The diffusion of oxygen through the plasma is considered a rate-limiting step in getting oxygen into the tissues.^8^ Fick’s Law predicts that if anatomical relationships remain constant, the change in the concentration (flux) of a substance is directly proportional to both the concentration gradient and the diffusivity. Diffusivity is the ease with which a compound moves due to Brownian motion through a medium. The uptake of oxygen by tissues declines with hypoxemia since the driving concentration gradient from the bloodstream to the tissues is reduced. This results in less oxygen delivered per unit time. Therapies that provide supplemental oxygen to patients provide increased tissue oxygen levels in accordance with Fick’s Law by increasing the concentration difference of oxygen between blood and tissue. Modifying the diffusivity of oxygen to improve end-organ delivery and to improve clinical outcomes has not been explored previously in COVID-19 since there has not been a drug available to directly modify diffusivity. Based on nonclinical and in vitro evidence, it is suggested that the primary mechanism by which TSC produces physiological effects is by transiently increasing hydrogen bonds between water molecules within the plasma component of blood. By enhancing this organizational matrix within plasma, wider and more direct pathways are created that allow oxygen to passively diffuse from high concentration areas to low concentration areas encountering less resistance.^9,10^ This increased diffusivity, or transfer efficiency, of oxygen to tissues in patients with hypoxemia may improve clinical outcomes.

This report describes the results from the lead-in phase. The lead-in phase was a multiple ascending dose study primarily investigating the safety and pharmacokinetics of TSC with multiple daily dosing not previously studied. Pre-defined clinical efficacy outcomes were also analyzed but no formal statistical analyses were conducted for efficacy; however, descriptive results for efficacy endpoints are presented. The WHO 9-point ordinal scale assessment was the first assessment of the subject’s clinical status each day, recording the worst score for the previous day. A reduction in the score derived from this scale is indicative of subject improvement. In the lead-in phase of the study, there was an observed reduction in the amount of time to improvement in WHO Ordinal Scale with increasing dose of TSC, ranging from a median of 11.5 days in the 0.25 mg/kg dose group to 7 days in the 1.5 mg/kg dose group. A total of 36% of subjects had a 1-point improvement in WHO Ordinal Scale to Day 7. The 1.0 and 1.5 mg/kg dose groups showed improvement in median pre-dose S/F ratio at Day 5 and later, not seen for 0.25 mg/kg and 0.50 mg/kg. The 1.5 mg/kg dose resulted in observed superior outcomes for multiple secondary clinical outcomes: time to 1-point WHO Ordinal Score improvement through day 29/discharge, 1-point improvement by day 7, days to return to room air, and hospital length of stay. Another indicator of subject improvement with TSC treatment included an increase in median SpO_2_/FiO_2_ (S/F) over time; these results are observational.

No new safety signals were identified as a result of this study. There were no TSC related TEAEs reported. Serious AEs and the death of 1 subject were assessed to be related to the underlying condition of SARS-CoV-2 infection. The results of the PK analyses in this study were consistent with other studies with TSC showing that increasing dose of TSC was associated with a decrease in clearance. There was no evidence that clearance was affected by covariates other than dose level.

These findings suggest that administration of TSC every 6 hours daily at doses up to 1.5 mg/kg for up to 15 days was well tolerated with a predictable pharmacokinetic profile independent of covariates aside from dose. Additionally, there were observed improvements in multiple clinical endpoints at the higher doses of TSC in the treatment of patients with COVID-19-related hypoxia.

## Data Availability

All data produced in the present study are available upon reasonable request to the authors

https://clinicaltrials.gov/ct2/show/NCT04573322

## DATA SHARING

A data sharing statement provided by the authors is available with the full text of this article.

## SUPPORTED BY

Supported by Diffusion Pharmaceuticals, Inc.

## FINANCIAL DISCLOSURE

Disclosure forms provided by the authors are available with the full text of this article.

## ACKNOWLEDGEMENT

We thank the study participants; their families; the investigational site members involved in this trial; the Trial Team; the members of the Data and Safety Monitoring Board; and our CRO partners.

## Supplemental Tables

**Table S 1.** Subject Demographics.

| Parameter | Category | 0.25 mg/kg<br>(N = 6) | 0.50 mg/kg<br>(N = 6) | 1.00 mg/kg<br>(N = 7*) | 1.50 mg/kg<br>(N = 6) | Total<br>(N = 25) |
| --- | --- | --- | --- | --- | --- | --- |
| Age (years) | N | 6 | 6 | 7 | 6 | 25 |
|  | Mean (SD) | 51.50 (18.87) | 60.83 (16.04) | 54.43 (8.16) | 57.50 (8.89) | 56.00 (13.16) |
|  | Median (IQR) | 50.00 (23) | 67.50 (14.5) | 52.00 (13.5) | 56.00 (9.5) | 55.00 (15) |
|  | Min - Max | 29.00 - 80.00 | 31.00 - 72.00 | 43.00 - 63.00 | 50.00 - 73.00 | 29.00 - 80.00 |
|  | < 65 Years, n (%) | 4 (66.7%) | 3 (50.0%) | 7 (100.0%) | 5 (83.3%) | 19 (76.0%) |
|  | >= 65 Years, n (%) | 2 (33.3%) | 3 (50.0%) | 0 (0.0%) | 1 (16.7%) | 6 (24.0%) |
| Sex | Female, n (%) | 3 (50.0%) | 2 (33.3%) | 4 (57.1%) | 4 (66.7%) | 13 (52.0%) |
|  | Male, n (%) | 3 (50.0%) | 4 (66.7%) | 3 (42.9%) | 2 (33.3%) | 12 (48.0%) |
| Race | White, n (%) | 6 (100.0%) | 6 (100.0%) | 7 (100.0%) | 6 (100.0%) | 25 (100.0%) |
| Weight (kg) | N | 6 | 6 | 7 | 6 | 25 |
|  | Mean (SD) | 79.00 (17.54) | 90.67 (13.84) | 89.57 (18.55) | 95.65 (20.92) | 88.76 (17.83) |
|  | Median (IQR) | 86.50 (16.5) | 87.50 (11) | 92.00 (18) | 95.50 (9.175) | 91.50 (13) |
|  | Min - Max | 47.00 - 92.00 | 75.00 - 115.00 | 64.00 - 117.00 | 64.00 - 129.00 | 47.00 - 129.00 |
| Height (cm) | N | 6 | 6 | 7 | 6 | 25 |
|  | Mean (SD) | 170.67 (5.35) | 168.17 (7.60) | 169.57 (10.77) | 168.33 (13.87) | 169.20 (9.39) |
|  | Median (IQR) | 168.50 (5.5) | 167.00 (9.75) | 170.00 (9.5) | 167.00 (18.25) | 170.00 (11) |
|  | Min - Max | 166.00 - 180.00 | 160.00 - 180.00 | 150.00 - 182.00 | 150.00 - 187.00 | 150.00 - 187.00 |
| BMI (kg/m<sup>2</sup>) | N | 6 | 6 | 7 | 6 | 25 |
|  | Mean (SD) | 27.08(5.84) | 31.90(2.46) | 30.88(3.86) | 33.64(5.79) | 30.87(4.98) |
|  | Median (IQR) | 28.23 (4.92) | 32.23 (1.0375) | 31.83 (4.265) | 32.81 (6.3975) | 31.14 (4.55) |
|  | Min - Max | 16.65 - 32.99 | 27.89 - 35.49 | 24.09 - 35.32 | 28.44 - 43.60 | 16.65 - 43.60 |

\* One subject received 1 dose on Day 1 and voluntarily withdrew
Source: Diffusion Pharmaceuticals, Inc. Clinical Study Report: 100-303, Table 14.1.2.1
Percentages are based on the Safety Population.

**Table S 2.** Summary of Time* (Days) to 1-Point Improvement in WHO Ordinal Severity Scale Through Day 29/Discharge.

| Statistics | 0.25 mg/kg<br>(N = 6) | 0.50 mg/kg<br>(N = 6) | 1.00 mg/kg<br>(N = 7) | 1.50 mg/kg<br>(N = 6) | Total<br>(N = 25) |
| --- | --- | --- | --- | --- | --- |
| N | 6 | 6 | 7 | 6 | 25 |
| Mean (SD) | 14.33 (11.54) | 13.67 (8.43) | 7.71 (4.31) | 7.50 (1.22) | 10.68 (7.63) |
| Median | 11.50 | 10.50 | 8.00 | 7.00 | 7.00 |
| Min - Max | 3.00 - 28.00 | 7.00 - 28.00 | 1.00 - 14.00 | 7.00 - 10.00 | 1.00 - 28.00 |

Source: Table 14.2.1.1
\*: Subjects who had no 1-point improvement in WHO ordinal severity scale during the study, the time (day) to 1-point improvement was imputed by last study day (Day 29/Discharge).

**Figure S 1.**
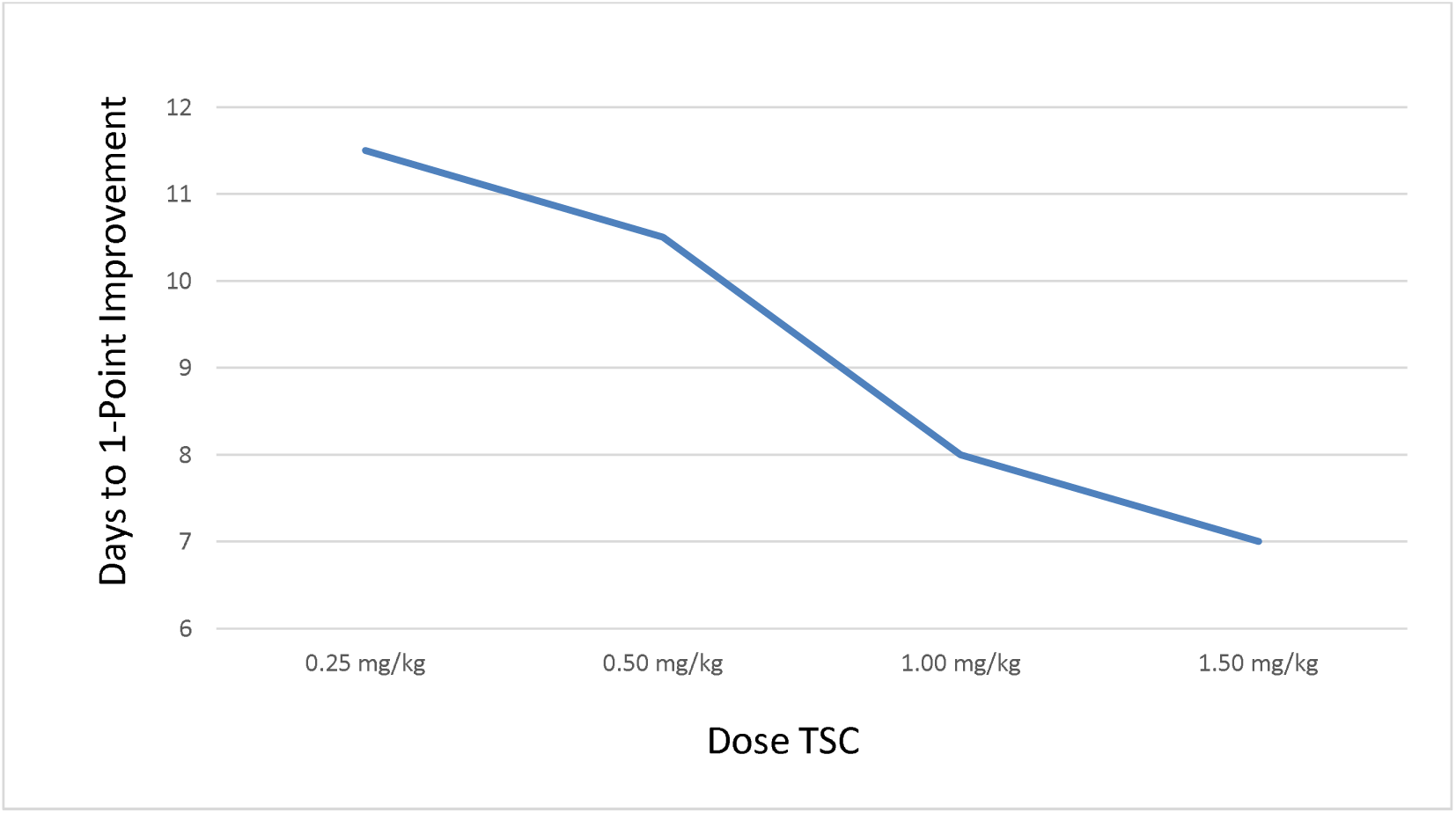
Change in Median Number of Days to 1-Point Improvement in WHO Ordinal Severity Scale.

**Table S 3.** Number and Percentage of Patients by WHO Ordinal Severity Scale Change from Baseline to Day 7.

| Category Change | 0.25 mg/kg<br>(N = 6) | 0.50 mg/kg<br>(N = 6) | 1.00 mg/kg<br>(N = 7) | 1.50 mg/kg<br>(N = 6) | Total<br>(N = 25) |
| --- | --- | --- | --- | --- | --- |
| 1 Point Worse from Day 1 to Day 7 | 0 (0.0%) | 1 (16.7%) | 0 (0.0%) | 0 (0.0%) | 1 (4.0%) |
| No Change from Day 1 to Day 7 | 3 (50.0%) | 3 (50.0%) | 5 (71.4%) | 1 (16.7%) | 12 (48.0%) |
| 1 Point Improvement from Day 1 to Day 7 | 1 (16.7%) | 2 (33.3%) | 1 (14.3%) | 5 (83.3%) | 9 (36.0%) |
| No Data Collected on Day 7 | 2 (33.3%) | 0 (0.0%) | 1 (14.3%) | 0 (0.0%) | 3 (12.0%) |

Source: Table 14.2.1.2

**Table S 4.**
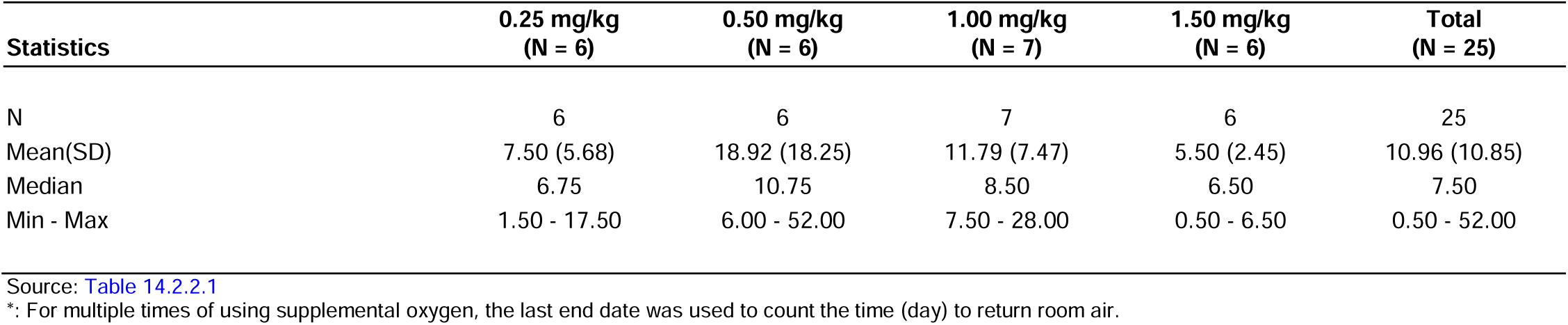
Summary of Time* (Days) to Return to Room Air by Treatment Group.

| Statistics | 0.25 mg/kg<br>(N = 6) | 0.50 mg/kg<br>(N = 6) | 1.00 mg/kg<br>(N = 7) | 1.50 mg/kg<br>(N = 6) | Total<br>(N = 25) |
| --- | --- | --- | --- | --- | --- |
| N | 6 | 6 | 7 | 6 | 25 |
| Mean(SD) | 7.50 (5.68) | 18.92 (18.25) | 11.79 (7.47) | 5.50 (2.45) | 10.96 (10.85) |
| Median | 6.75 | 10.75 | 8.50 | 6.50 | 7.50 |
| Min - Max | 1.50 - 17.50 | 6.00 - 52.00 | 7.50 - 28.00 | 0.50 - 6.50 | 0.50 - 52.00 |

Source: Table 14.2.2.1
\*: For multiple times of using supplemental oxygen, the last end date was used to count the time (day) to return room air.

**Table S 5.** Overall Summary of Subjects with TEAE during Lead-In Phase Safety Population.

|  | **0.25 mg/kg**<br>**(N = 6)** | **0.50 mg/kg**<br>**(N = 6)** | **1.00 mg/kg**<br>**(N = 7)** | **1.50 mg/kg**<br>**(N = 6)** | **Total**<br>**(N = 25)** |
| --- | --- | --- | --- | --- | --- |
| Patients with any TEAE | 3 (50.0%) | 4 (66.7%) | 5 (71.4%) | 2 (33.3%) | 14 (56.0%) |
| Patients with any Serious TEAE | 1 (16.7%) | 1 (16.7%) | 0 (0.0%) | 0 (0.0%) | 2 (8.0%) |
| Patients with any TEAE of DLT | 0 (0.0%) | 0 (0.0%) | 0 (0.0%) | 0 (0.0%) | 0 (0.0%) |
| Patients with any TEAE resulting in death | 1 (16.7%) | 0 (0.0%) | 0 (0.0%) | 0 (0.0%) | 1 (4.0%) |
| Patients with any TEAE leading to study drug discontinuation | 0 (0.0%) | 1 (16.7%) | 0 (0.0%) | 0 (0.0%) | 1 (4.0%) |
| Patients with any TEAE leading to study drug interruption | 0 (0.0%) | 0 (0.0%) | 0 (0.0%) | 0 (0.0%) | 0 (0.0%) |
| Patients with any TEAE with CTCAE Grade 3 or 4 | 1 (16.7%) | 3 (50.0%) | 0 (0.0%) | 1 (16.7%) | 5 (20.0%) |

Source: Table 14.3.1.1
% is based on the total numbers of Safety Population,
TEAE = Treatment-Emergent Adverse Events, TDL = Dose Limiting Toxicity

**Table S 6.**
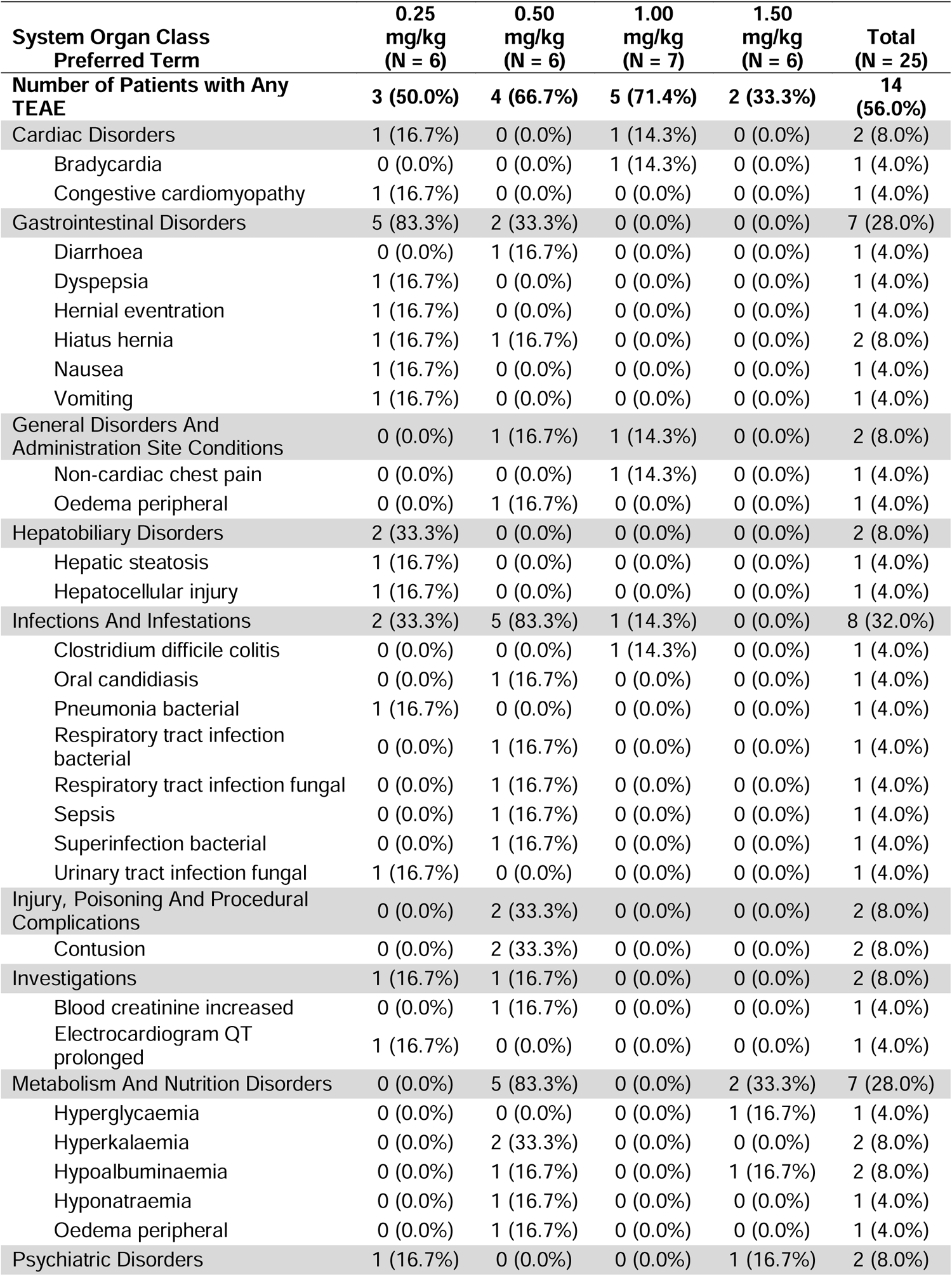
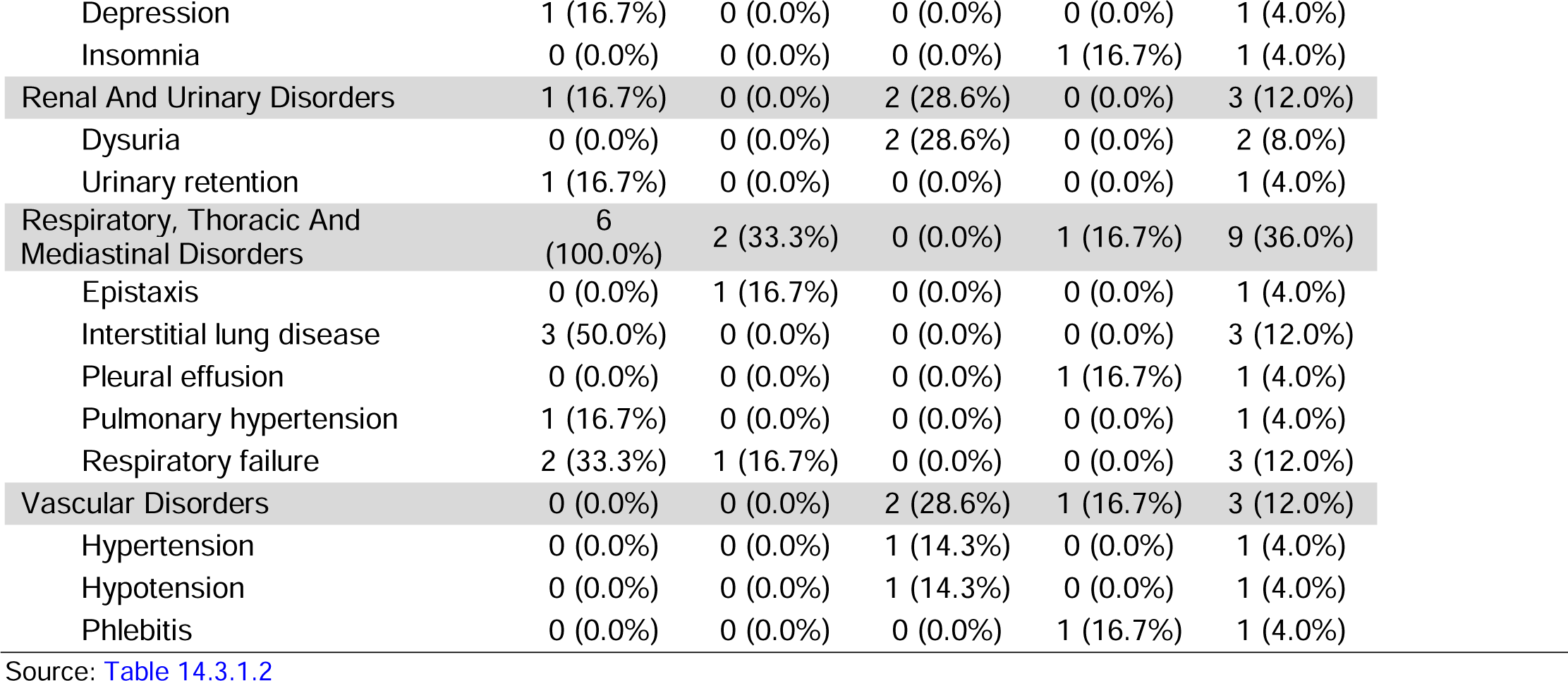
TEAEs Reported in ≥2 Subjects per SOC During the Lead-in Phase - Safety Population.

**Figure S 2.**
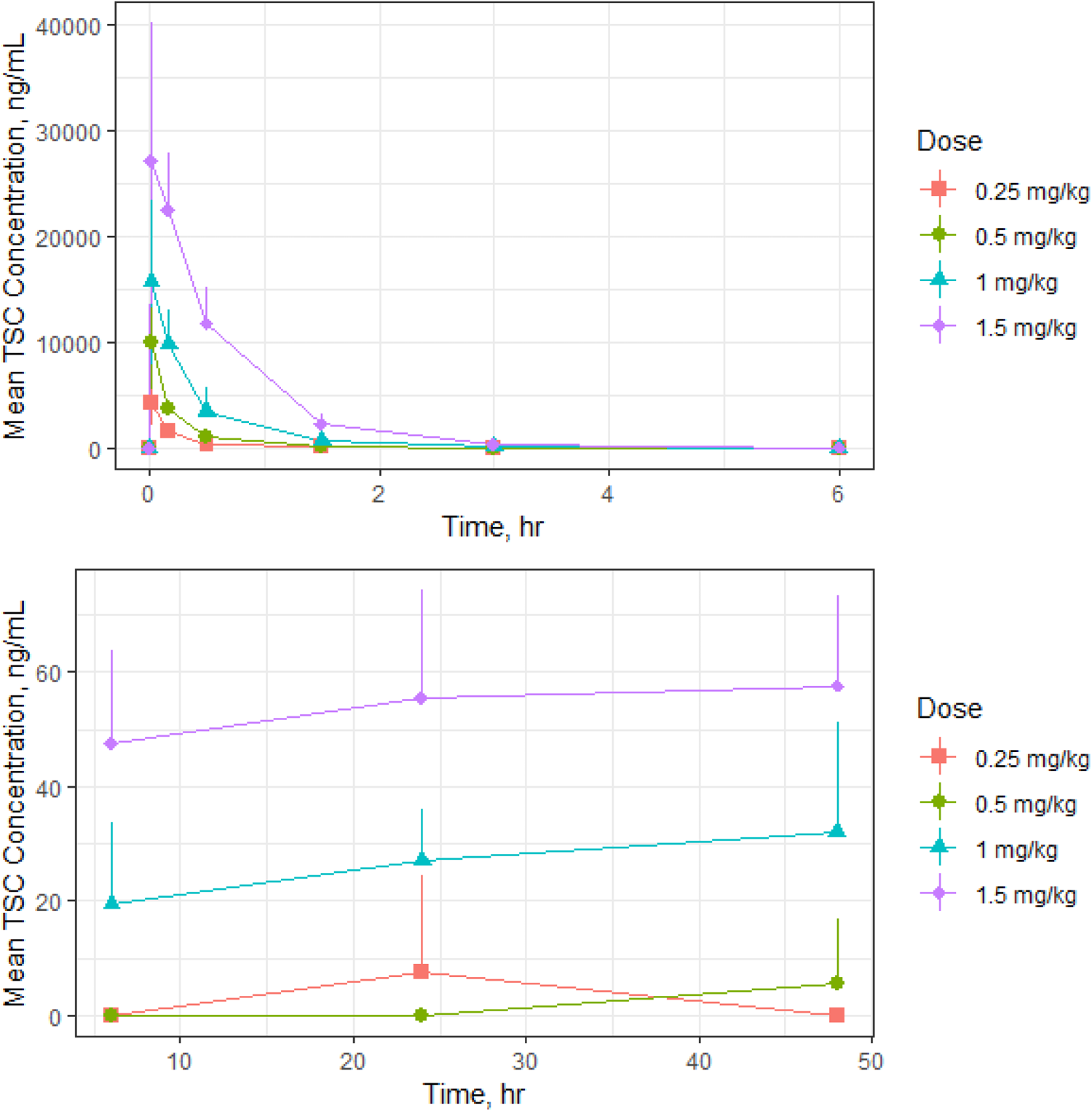
Mean (+SD) TSC Plasma Concentration-Time Profiles by Dose Cohort Following Multiple IV Administration of TSC Every Six Hours, Linear Scale.

**Table S 7.** Summary of Pharmacokinetic Parameters of TSC Following IV Bolus Administration of TSC by Dose Cohort of Patients with SARS-CoV-2 Infection and Hypoxemia in Study 100-301.

| Parameter | Dose |  |  |  |
| --- | --- | --- | --- | --- |
|  | 0.25 mg/kg<br>(N=6) | 0.5 mg/kg<br>(N=6) | 1 mg/kg<br>(N=7) | 1.5 mg/kg<br>(N=6) |
| C<sub>max</sub> (± SD) ng/mL | 4197 ± 1295 | 10072 ± 3188* | 17829 ± 3519 | 28783 ± 9765 |
| T<sub>max</sub> h | 0.0167 | 0.0167* | 0.0167 | 0.0919 |
| AUC<sub>0-t</sub> (± SD) h*ng/mL | 910 ± 374 | 2240 ± 335* | 6293 ± 2564 | 16682 ± 4831 |
| AUC<sub>0-∞</sub> (± SD) h*ng/mL | 934 ± 374 | 2264 ± 330* | 6332 ± 2563 | 16756 ± 4844 |
| t<sub>1/2z</sub> (± SD) h | 0.53 ± 0.156 | 0.604 ± 0.0535* | 0.929 ± 0.317 | 1.07 ± 0.118 |
| CL (± SD) mL/min/kg | 5.38 ± 2.96 | 3.74 ± 0.525* | 2.97 ± 1.02 | 1.62 ± 0.563 |
| V<sub>z</sub> (± SD) L/kg | 0.225 ± 0.0776 | 0.197 ± 0.0415* | 0.238 ± 0.111 | 0.151 ± 0.0552) |

Source: PK Study Report Protocol number: 100-303
AUC<sub>0-∞</sub>, area under the concentration curve from time zero to infinity; AUC<sub>0-t</sub>, area under the concentration vs time curve from time zero to tau; C<sub>max</sub>, maximum concentration; CL, clearance; SD, standard deviation; t<sub>1/2z</sub>, terminal elimination half-life; T<sub>max</sub>, time to maximum concentration V<sub>z</sub>, volume in terminal state
\*n=5 Samples from 1 subject were excluded from the analysis because PK profile was not recognized as a PK profile after an IV bolus dose

**Table S 8.** Change in Median SpO_2_/FiO_2_ Ratio Over Time.

|  | 0.25<br>mg/kg | 0.50<br>mg/kg | 1.00<br>mg/kg | 1.50<br>mg/kg | Total |
| --- | --- | --- | --- | --- | --- |
| Baseline | 284.55 | 127.35 | 176.4 | 245 | 217.8 |
| Day 2 | 302.55 | 128 | 207 | 263.95 | 221.35 |
| Day 3 | 255.85 | 132.3 | 232.5 | 277.55 | 242.5 |
| Day 4 | 294.45 | 130.2 | 289.05 | 330 | 275 |
| Day 5 | 241.95 | 132.25 | 316.55 | 369.15 | 304.7 |
| Day 6 | 301.9 | 239.65 | 326.7 | 461.9 | 330 |
| Day 7 | 219.6 | 132 | 405.35 | 461.9 | 341.8 |
| Day 8 | 219.7 | 142 | 410.15 | 461.9 | 331.5 |
| Day 9 | 291.7 | 158.1 | 471.4 | 457.1 | 452.4 |
| Day 10 | 122.7 | 158.5 | 469.05 | 466.7 | 324.25 |

**Figure S 3.**
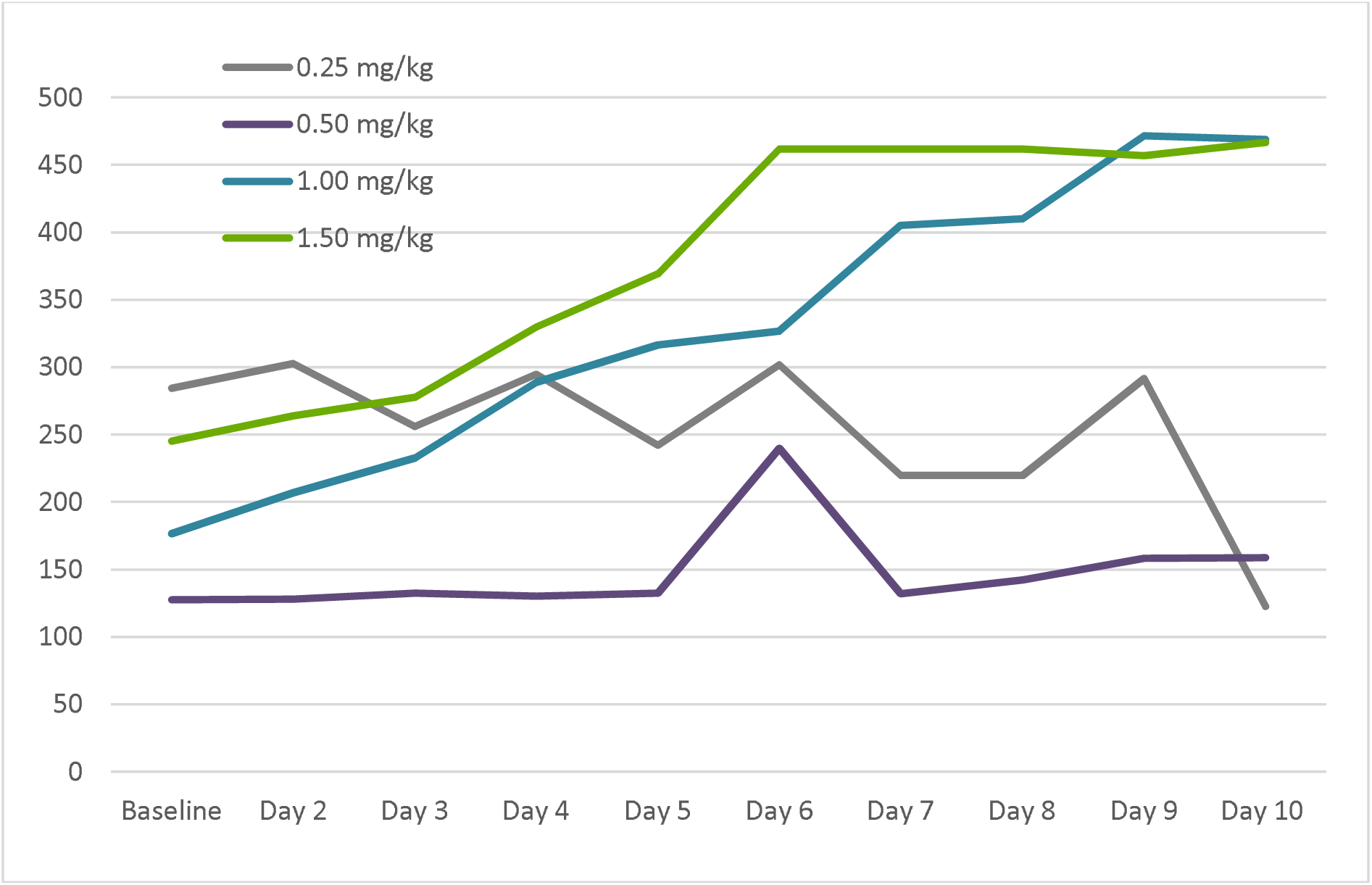
Summary of Oxygen Saturation (via Pulse Oximetry) Results by Study Day: Median SpO_2_/FiO_2_ Ratio.

### Appendix A Inclusion/Exclusion Criteria

#### Inclusion Criteria

1. Hospitalized subjects with confirmed SARS-CoV-2 infection and hypoxemia, defined as SpO2 < 94% on room air or requiring supplemental oxygen (inclusive of nasal cannula, high flow oxygen, non-invasive ventilation, and mechanical ventilation)
2. Laboratory-confirmed SARS-CoV-2 infection as determined by rtPCR, or other commercial or public health assay in any specimen within 7 days prior to enrollment.
3. WHO ordinal scale score of 3 through 7 (exclusive of ECMO) at baseline
4. Male or non-pregnant female adult ≥18 years of age at time of enrollment.
5. Subject (or legally authorized representative) provides written informed consent prior to initiation of any study procedures.
6. Understands and agrees to comply with planned study procedures.
7. Illness of any duration
8. Women of childbearing potential must have a negative blood pregnancy test at the screening/baseline visit and agree to use two forms of birth control through 30 days after the last dose of study drug.

#### Exclusion Criteria

1. Receiving extracorporeal membrane oxygenation (ECMO) at baseline
2. Severe organ dysfunction (SOFA score > 10) at enrollment
3. Patient or LAR unable to provide written informed consent
4. ALT/AST > 3 times the upper limit of normal or serum bilirubin > 1.5 times the upper limit of normal
5. Estimated glomerular filtration rate (eGFR) by Modification of Diet in Renal Disease (MDRD) formula < 30 mL/min/1.73 m2 or on dialysis
6. Pregnancy or breast feeding.
7. Anticipated transfer to another hospital which is not a study site within 72 hours.
8. Allergy to any study medication
9. Patient not expected to survive >24 hours, or likely to be discharged <24 hours per PI discretion.

## Notes

### Competing Interest Statement

The authors have declared no competing interest.

### Clinical Trial

NCT04573322

### Funding Statement

This study was funded by Diffusion Pharmaceuticals.

### Author Declarations

National Agency for Medicines and Medical Devices of Romania Medicinal Products for Human Use gave ethical approval for this work. 48 Av Sanatescu Street, District 1 011478, Bucharest, Romania

